# Risk Factors for Mortality of COVID-19 Patients

**DOI:** 10.1101/2020.07.02.20145375

**Authors:** Ouail Ouchetto, Asmaa Drissi Bourhanbour

## Abstract

**Background:** Lethality rates of COVID-19 are so different between countries and continents. This lethality seems to be very low in Africa and Asia, but exceedingly high in western Europe and North America. Many factors could have a role in this disparity such as comorbidities. Advanced age, obesity, cardiovascular disease, diabetes and cancer were the most frequently cited in the reported COVID-19 data. The main objective was to analyze and evaluate the association between the COVID-19 mortality and the mentioned factors in 164 countries.

**Methods:** The Data of COVID-19 deaths, latitude degrees, population age distribution, cardiovascular diseases, obesity, diabetes and cancer were extracted from different online sources. For the statistical analysis, we used Spearman to measure the correlation coefficient between numbers of deaths and the mentioned factors until June 29, 2020.

**Results:** The correlation between COVID-19 mortality and latitude, high age, obesity, CVD and number of cancer patients per 100,000 is significant at 0.01 level with r = 0.489, r=0.511, r=0.489, r=0.561 and r=0.536 respectively. The correlation between the number of deaths and diabetes is less strong than the previous ones, and the correlation coefficient is r= 0.154.

**Conclusion:** The great lethality of COVID-19 in western Europe and North America can be explained in part by the highest of age, cancer and CVD percentage in these regions. It seems also plausible that the increased obesity in the USA and vitamin D deficiency in Europe may contribute to increasing the number of COVID-19 deaths.

## Introduction

Coronavirus disease 2019 (COVID-19) is an infectious disease caused by severe acute respiratory syndrome coronavirus 2 (SARS-CoV-2), which has spread rapidly through China and the rest of the world. SARS-CoV-2 shares some characteristics with two other coronaviruses: severe acute respiratory syndrome coronavirus (SARS-CoV) appeared in 2003 and Middle East respiratory syndrome (MERS-CoV) appeared in 2012. COVID-19 had affected over 10 million people and killed more than 500,000 in over 200 countries worldwide as of 29 June 2020 [1,2]. According to the clinical data, some patients with COVID-19 developed acute respiratory distress syndrome and part of them worsened in a short period and died of multiple organ failure.

Researchers have emphasized that patients at risk for severe acute respiratory syndrome coronavirus-2 have been characterized as having advanced age or preexisting diseases, like obesity, cardiovascular disease, diabetes, cancer or chronic respiratory disease (A1). In [3], the presented data support that diabetes should be considered as a risk factor for a rapid progression and bad prognosis of COVID-19 and in case of rapid deterioration, more intensive attention should be paid to diabetic patients. In addition, data from Italy showed more than two-thirds of those who died by severe acute respiratory syndrome coronavirus 2 (SARS-CoV-2) had diabetes [4]. Since obesity has been shown to increase vulnerability to infections, it may be a risk factor for COVID□19□ related mortality [5]. The Seattle region was among the first to report body mass index (BMI) data, and a sample has showed that 85% of patients with obesity required mechanical ventilation and 62% of patients with obesity died. These proportions are greater than those in patients without obesity, in which 64% required mechanical ventilation and 36% died [6]. Cardiovascular disease is mentioned as one of the most common comorbidities. In Chinese report from 138 hospitalized COVID□19 patients, 14.5% had cardiovascular disease [7]. Another study on 99 hospitalized patients in China showed that 40% of the cohort had cardiovascular or cerebrovascular disease [8]. Cancer can be also a risk factor of COVID-19. In fact, a study of small sample size of SARS-CoV-2 patients reported a higher risk of severe events in patients with cancer when compared with patients without cancer [9]. A multicenter study including 105 patients with cancer and 536 noncancer patients infected with COVID-19 confirmed also that the COVID-19 patients with cancer had higher risks in all severe outcomes. Patients with hematologic cancer, lung cancer, or with metastatic cancer (stage IV) had the highest frequency of severe events [10].

The lethality rates of COVID-19 are very different between countries and continents. This lethality seems to be very low in Africa, Asia and in some South America countries, and high in western Europe and North America. In this work, we analyze and evaluate the association between the deaths number of COVID-19 and of the mentioned risk factors in 164 countries.

## Method

The Data of COVID-19 of the studied countries were mainly obtained from Worldometers [1] and European Centre for Disease Prevention and Control [2], as of 29 June 2020. The percentage of obese adults were gathered from the procon.org organization [11] and the percentage of the diabetic population aged between 20 to 79 years were collected from the Index Mundi database [12]. In the other side, the standardized incidence rates of all cancers (per 100,000) were obtained from the Global cancer observatory [13] and the percentages of cardiovascular diseases were extracted from the Global Health Data Exchange (GHDx) database [14]. Finally, the percentage of the population having more than 55 years and latitude degrees are available from [15, 16].

This study involves 164 countries around the world. Indeed, we have included all countries whose data on COVID-19 mortality, population age distribution, CVD, obesity, diabetes and cancer are available (see Appendix). As the data were not normally distributed, we used the Spearman test for correlation analysis between the numbers of deaths of COVID-19 per 1 million population in the studied countries and the mentioned factors. Spearman’s correlation assesses monotonic relationships whether linear or not [17], and it doesn’t depend on the sample sizes and distributions of the variables. The Spearman’s correlation between two variables is low when observations have a dissimilar rank between the two variables, and high when observations have a similar rank between the two variables. In the case of no repeated data values, a perfect correlation of +1 or −1 is obtained when each of the variables is a perfect monotone function of the other.

## Results

Fig. 1 (a) represents the COVID-19 mortality per million as a function of the latitude degrees. It can be seen that all countries that locate below 35 degrees North have relatively low mortality and they have low than 300 deaths per million population. The correlation between mortality and latitude is r = 0.489 (see Table 1). The distribution of COVID-19 mortality versus high age (55 years and over) is plotted in Fig. 1 (b). The number of deaths correlates positively and significantly with high age (r = 0.511). Indeed, the countries with a high number of aged people have the highest deaths number.

**Table 1:** Correlation coefficients between COVID-19 mortality and high age, obesity, diabetes, cardiovascular diseases (CVD) and cancer.

| Correlations |  |  |  |  |  |  |  |
| --- | --- | --- | --- | --- | --- | --- | --- |
|  |  | Latitude | Age (55 years & over) | Diabetes | Obesity | Cardiovascular Diseases | Cancer |
| Mortality per million | Correlation Coefficient | 0.489** | 0.511** | 0.154* | 0.489** | 0.561** | 0.536** |
|  | Sig. (2-tailed) | 0.000 | 0.000 | 0.048 | 0.000 | 0.000 | 0.000 |
|  | N | 164 | 164 | 164 | 164 | 164 | 164 |
| **. Correlation is significant at the 0.01 level (2-tailed). |  |  |  |  |  |  |  |
| *. Correlation is significant at the 0.05 level (2-tailed). |  |  |  |  |  |  |  |

**Figure 1:**
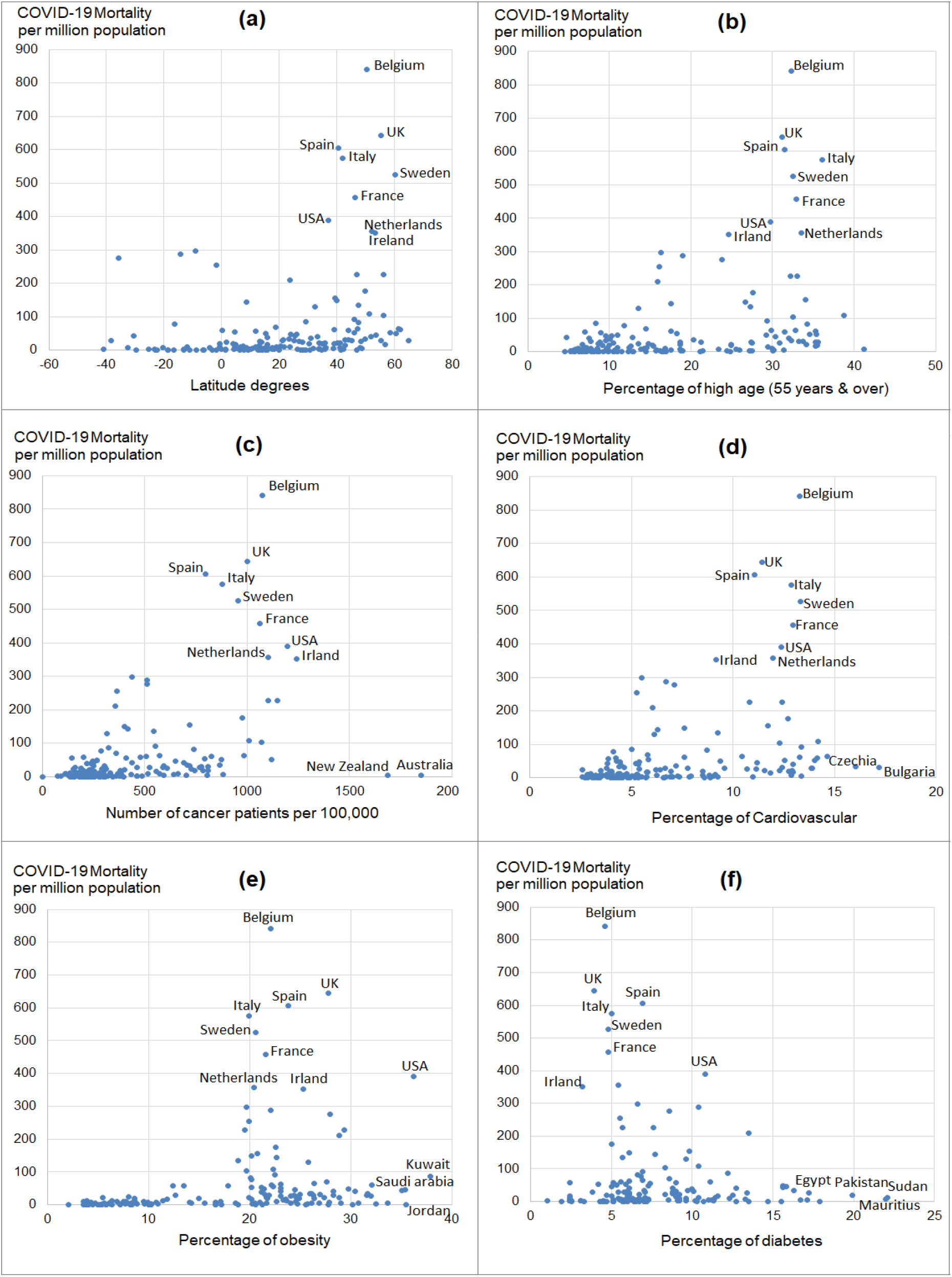
Association between COVID-19 mortality per million by country as a function of latitude, percentage of high age (55 years and over), number of cancer patients per 100,000, percentage of cardiovascular diseases, percentage of obesity and percentage of diabetes.

Except for Australia and New Zealand, the countries with a high number of cancer patients have the highest deaths number (Fig. 1(c)). The correlation is positive between the number of deaths and number of cancer patients per 100,000 (r = 0.591). In the other side, the association between the deaths number and CVD percentage is strong and the correlation coefficient is r = 0,56 (at 0.01 level). The distribution of CVD is presented in (Fig. 1(d)).

Fig. 1 (e) and Fig. 1 (g) represent the number of deaths versus the percentage of obesity and diabetes, respectively. The association between obesity and the number of deaths is significant with r=0.489. However, the association between diabetes and the number of deaths is less strong than the previous ones with r= 0,154.

## Discussion

The lethality rates of COVID-19 are very different between countries and continents. This lethality seems to be high in western Europe and North America compared to others countries. This North-South difference may be due to several factors. Firstly, the correlation between the number of mortality and latitude degrees can be explained by the vitamin D deficiency, which associated with increased risk and greater severity of COVID-19 [18]. Indeed, thirty-five degrees North also happens to be the latitude above which people do not receive sufficient sunlight to maintain adequate vitamin D levels during winter. In Europe, the prevalence of hypovitaminosis D is high, but it is relatively uncommon in Nordic countries, probably due to the widespread use of supplements [19], which can explain that the mortality is relatively low in these countries. This correlation can be also explained by the existing of the high prevalence of the elderly population in western Europe and North America. Older people are more likely to develop risk factors and severe health consequences from COVID-19 [20].

Secondly, the percentage of CVD is high in western Europe and North of America, which explains the significant association between mortality and the percentage of CVD. Indeed, CVD patients are more likely to be infected due to their deteriorated heart function. In fact, pre-existing CVD heightens the vulnerability to develop COVID-19 and to have more severe disease with worse clinical outcomes and prognosis [21].

The incidence rate of all cancers remains also high in western Europe and North of America, which provides a significant correlation with the mortality rate. Cancer patients are at high risk for infections due to coexisting chronic diseases, overall weakened health status, and systemic immunosuppressive states caused by both cancer and anticancer treatments [22]. As a consequence, cancer patients infected by the SARS-CoV-2 coronavirus may experience more difficult outcomes than other populations.

The correlation between the mortality of COVID-19 and obesity was significant. Even if the obesity prevalence in Europe is not the highest in the world, but it remains high in the North of America. In fact, obesity is characterized by chronic inflammation associated with a decreased immune system, leading to susceptibility to infection [23]. Additionally, patients with obesity show a restrictive breathing pattern and reduced lung volume. This evidence suggests that obesity might act as an independent risk factor for a poor disease progression of COVID-19.

Diabetes mellitus is one of the most common chronic diseases and one of the leading causes of morbidity worldwide [24]. Several studies have demonstrated that diabetics have high susceptibility to some infectious diseases, like Mycobacterium tuberculosis and Staphylococcus aureus [25]. However, the correlation between diabetes and mortality is not significant and this is due to the lowest of prevalence of diabetes in western Europe and north America countries.

## Conclusion

The computed correlation coefficients between the risk factors and the mortality numbers of COVID-19 show that the great lethality of COVID-19 in western Europe and North America can be explained in part by the highest of age, cancer and CVD prevalence in these regions. It seems also plausible that the increased obesity in the USA and vitamin D deficiency in Europe may contribute to increasing the number of COVID-19 deaths. In the other side, the correlation between diabetes and the number of COVID-19 deaths is not very significant.

## Data Availability

Links of the used data are in reference of the manuscript

## Acknowledgments

None.

## Ethical Approval

Approval was not required

## Appendix

**Table:** Worldwide mortality rate of COVID-19, latitude degrees, percentage of age (55 years and over), obesity, diabetes, cardiovascular disease (CVD) and cancer (per 100,000).

| Country | Deaths/<br>1 Million<br>population | Latitude<br>Degrees | Cancer/<br>(100.000) | Diabetes<br>(%) | Obesity<br>(%) | CVD<br>(%) | Age 55<br>years &<br>over (%) |
| --- | --- | --- | --- | --- | --- | --- | --- |
| Belgium | 841 | 50.50 | 1076 | 4.6 | 22.1 | 13.27 | 32.35 |
| UK | 644 | 55.38 | 1001.9 | 3.9 | 27.8 | 11.42 | 31.21 |
| Spain | 606 | 40.46 | 798.2 | 6.9 | 23.8 | 11.06 | 31.48 |
| Italy | 575 | 41.87 | 878.4 | 5 | 19.9 | 12.87 | 36.08 |
| Sweden | 526 | 60.13 | 956.8 | 4.8 | 20.6 | 13.33 | 32.49 |
| France | 457 | 46.23 | 1060.7 | 4.8 | 21.6 | 12.95 | 32.93 |
| United States | 390 | 37.09 | 1195.7 | 10.8 | 36.2 | 12.37 | 29.71 |
| Netherlands | 357 | 52.13 | 1103.4 | 5.4 | 20.4 | 11.99 | 33.51 |
| Ireland | 352 | 53.41 | 1240.5 | 3.2 | 25.3 | 9.18 | 24.59 |
| Chile | 298 | -35.68 | 512.1 | 8.6 | 28 | 7.11 | 23.79 |
| Peru | 288 | -9.19 | 438.8 | 6.6 | 19.7 | 5.52 | 16.33 |
| Brazil | 277 | -14.24 | 513.3 | 10.4 | 22.1 | 6.7 | 18.99 |
| Ecuador | 255 | -1.83 | 362.9 | 5.5 | 19.9 | 5.25 | 16.07 |
| Canada | 227 | 56.13 | 1148.3 | 7.6 | 29.4 | 12.44 | 33.06 |
| Switzerland | 227 | 46.82 | 1102.9 | 5.7 | 19.5 | 10.83 | 32.21 |
| Mexico | 210 | 23.63 | 354.2 | 13.5 | 28.9 | 6.03 | 15.96 |
| Luxembourg | 176 | 49.82 | 975.6 | 5 | 22.6 | 12.73 | 27.56 |
| Portugal | 155 | 39.40 | 719.2 | 9.8 | 20.8 | 11.74 | 34 |
| Armenia | 149 | 40.07 | 402.2 | 6.1 | 20.2 | 7.62 | 26.68 |
| Panama | 144 | 8.54 | 418.3 | 7.7 | 22.7 | 6.29 | 17.55 |
| Moldova | 135 | 47.41 | 545.5 | 5.7 | 18.9 | 9.25 | 27.29 |
| Iran | 129 | 32.43 | 314.2 | 9.6 | 25.8 | 6.15 | 13.59 |
| Germany | 108 | 51.17 | 1010.9 | 10.4 | 22.3 | 14.21 | 38.73 |
| Denmark | 104 | 56.26 | 1068.9 | 8.3 | 19.7 | 12.29 | 32.54 |
| Bolivia | 92 | -16.29 | 285.4 | 6.8 | 20.2 | 4.13 | 11.77 |
| Romania | 86 | 45.94 | 552.6 | 6.9 | 22.5 | 13.37 | 29.31 |
| Kuwait | 83 | 29.31 | 321.8 | 12.2 | 37.9 | 5.01 | 8.35 |
| Austria | 78 | 47.52 | 737.9 | 6.6 | 20.1 | 8.73 | 34.28 |
| Dominican Republic | 69 | 18.74 | 361.5 | 8.6 | 27.6 | 5.82 | 14.46 |
| Russia | 64 | 61.52 | 571.3 | 6.1 | 23.1 | 10.46 | 29.84 |
| Colombia | 63 | 4.57 | 407.8 | 7.4 | 22.3 | 5.84 | 18.32 |
| Hungary | 61 | 47.16 | 984 | 6.9 | 26.4 | 14.63 | 32.86 |
| Turkey | 61 | 38.96 | 508.3 | 11.1 | 32.1 | 7.63 | 17.6 |
| Finland | 59 | 61.92 | 825.6 | 5.6 | 22.2 | 13.28 | 35.28 |
| Sao Tome | 59 | 0.19 | 199.2 | 2.4 | 12.4 | 4.23 | 7.04 |
| Bosnia & Herzegovina | 57 | 43.92 | 479.2 | 9 | 17.9 | 14.17 | 31.46 |
| Djibouti | 55 | 11.83 | 143.7 | 5.1 | 13.5 | 3.87 | 8.98 |
| Slovenia | 53 | 46.15 | 875.6 | 5.9 | 20.2 | 14.01 | 35.42 |
| Estonia | 52 | 58.60 | 794.8 | 4.2 | 21.2 | 12.54 | 34.58 |
| Bahrain | 51 | 25.93 | 265.9 | 15.6 | 29.8 | 5.39 | 10.25 |
| Honduras | 49 | 15.20 | 264.7 | 7.3 | 21.4 | 4.27 | 10.98 |
| Iraq | 48 | 33.22 | 219.1 | 8.8 | 30.4 | 5.34 | 7.56 |
| Saudi Arabia | 47 | 23.89 | 246.3 | 15.8 | 35.4 | 4.45 | 9.58 |
| Norway | 46 | 60.47 | 1120.3 | 5.3 | 23.1 | 9.39 | 29.27 |
| South Africa | 43 | -30.56 | 436.2 | 12.7 | 28.3 | 5.21 | 12.89 |
| Guatemala | 42 | 15.78 | 238.9 | 10 | 21.2 | 4.47 | 10.11 |
| Belarus | 41 | 53.71 | 649 | 5 | 24.5 | 11.18 | 30.38 |
| Qatar | 40 | 25.35 | 250.3 | 15.6 | 35.1 | 3.82 | 4.72 |
| Poland | 39 | 51.92 | 691.5 | 6.1 | 23.1 | 12.96 | 32.04 |
| Israel | 35 | 31.05 | 706.4 | 9.7 | 26.1 | 7.19 | 20.36 |
| Oman | 34 | 21.51 | 250 | 10.1 | 27 | 4.64 | 7.7 |
| Czechia | 33 | 49.82 | 868.2 | 7 | 26 | 16.03 | 32.35 |
| Bulgaria | 32 | 42.73 | 591.8 | 6 | 25 | 17.2 | 33.21 |
| Serbia | 32 | 44.02 | 721.2 | 9 | 21.5 | 12.43 | 33.7 |
| UAE | 32 | 23.42 | 303.1 | 16.3 | 31.7 | 4.39 | 9.58 |
| Iceland | 29 | 64.96 | 810.8 | 5.8 | 21.9 | 9.58 | 27.41 |
| Lithuania | 29 | 55.17 | 775.5 | 3.8 | 26.3 | 13.87 | 35.55 |
| Argentina | 28 | -38.42 | 587.1 | 5.9 | 28.3 | 7.76 | 21.2 |
| Bahamas | 28 | 25.03 | 462.5 | 8.8 | 31.6 | 6.65 | 18.71 |
| Egypt | 28 | 26.82 | 308.3 | 17.2 | 32 | 5.37 | 10.52 |
| Mauritania | 28 | 21.01 | 161.8 | 7.1 | 12.7 | 3.77 | 8.82 |
| El Salvador | 27 | 13.79 | 290.3 | 8.8 | 24.6 | 6.22 | 14.83 |
| Ukraine | 27 | 48.38 | 524.5 | 6.1 | 24.1 | 11.13 | 30.9 |
| Croatia | 26 | 45.10 | 769.8 | 5.4 | 24.4 | 13.92 | 35.3 |
| Cape Verde | 25 | 16.00 | 254.2 | 2.4 | 11.8 | 5.81 | 12.6 |
| Barbados | 24 | 13.19 | 607.9 | 13.4 | 23.1 | 10.7 | 27.48 |
| Equatorial Guinea | 23 | 1.65 | 189.2 | 6 | 8 | 2.6 | 8.61 |
| Albania | 22 | 41.15 | 401.1 | 9 | 21.7 | 12.31 | 24.97 |
| Suriname | 22 | 3.92 | 381 | 12.5 | 26.4 | 6.95 | 15.33 |
| Algeria | 21 | 28.03 | 312.6 | 6.7 | 27.4 | 6.4 | 13.58 |
| Azerbaijan | 21 | 40.14 | 228.1 | 6.1 | 19.9 | 12.91 | 18.7 |
| Malta | 20 | 35.94 | 729.5 | 8.3 | 28.9 | 11.57 | 34.18 |
| Afghanistan | 19 | 33.94 | 151 | 9.2 | 5.5 | 3.48 | 6.69 |
| Gabon | 19 | -0.80 | 238.4 | 6 | 15 | 4.05 | 9.17 |
| Montenegro | 19 | 42.71 | 555.6 | 9 | 23.3 | 11.84 | 29.43 |
| Pakistan | 19 | 30.38 | 200.3 | 19.9 | 8.6 | 3.94 | 9.99 |
| Greece | 18 | 39.07 | 783.8 | 4.7 | 24.9 | 12.77 | 35.53 |
| Cyprus | 16 | 35.13 | 743.2 | 9 | 21.8 | 8.51 | 24.89 |
| Latvia | 16 | 56.88 | 808.6 | 5 | 23.6 | 12.94 | 35.27 |
| Guyana | 15 | 4.86 | 219.4 | 11.6 | 20.2 | 5.75 | 15.38 |
| India | 13 | 20.59 | 173.5 | 10.4 | 3.9 | 4.37 | 14.63 |
| Sudan | 13 | 12.86 | 158.4 | 22.1 | 8.6 | 4.05 | 7.16 |
| Cameroon | 12 | 7.37 | 180.5 | 6 | 11.4 | 3.02 | 6.98 |
| Guinea-Bissau | 12 | 11.80 | 135.5 | 2.4 | 9.5 | 2.83 | 6.2 |
| Philippines | 12 | 12.88 | 330.3 | 7.1 | 6.4 | 4.44 | 11.04 |
| Bangladesh | 11 | 23.68 | 168.3 | 9.2 | 3.6 | 5.29 | 14.23 |
| Indonesia | 11 | -0.79 | 298 | 6.3 | 6.9 | 5.25 | 16.81 |
| Nicaragua | 11 | 12.87 | 277.5 | 11.4 | 23.7 | 4.78 | 12.45 |
| Central African Republic | 10 | 6.61 | 110.8 | 6 | 7.5 | 2.93 | 7.67 |
| Kazakhstan | 10 | 48.02 | 384.4 | 6.1 | 21 | 6.27 | 18.68 |
| Yemen | 10 | 15.55 | 115.4 | 5.4 | 17.1 | 3.77 | 6.8 |
| Haiti | 9 | 18.97 | 207.3 | 6.7 | 22.7 | 4.3 | 9.63 |
| Kyrgyzstan | 9 | 41.20 | 237.9 | 6.1 | 16.6 | 4.65 | 13.89 |
| Comoros | 8 | -11.88 | 166.3 | 12.3 | 7.8 | 4.4 | 8.57 |
| Cuba | 8 | 21.52 | 516.8 | 9.6 | 24.6 | 9.12 | 29.91 |
| Japan | 8 | 36.20 | 662.4 | 5.6 | 4.3 | 12.88 | 41.24 |
| Mauritius | 8 | -20.35 | 367.6 | 22 | 10.8 | 8.53 | 23.39 |
| Sierra Leone | 8 | 8.46 | 141 | 2.4 | 8.7 | 3.56 | 7.59 |
| Uruguay | 8 | -32.52 | 643.2 | 7.3 | 27.9 | 8.84 | 25.5 |
| Brunei | 7 | 4.54 | 572.2 | 13.3 | 14.1 | 4.39 | 14.24 |
| Congo democratic | 7 | -4.04 | 153 | 6 | 9.6 | 3.47 | 5.83 |
| Liberia | 7 | 6.43 | 142.9 | 2.4 | 9.9 | 3.24 | 6.29 |
| Senegal | 7 | 14.50 | 189.5 | 2.4 | 8.8 | 3.52 | 7.31 |
| Mali | 6 | 17.57 | 202.3 | 2.4 | 8.6 | 2.9 | 6.7 |
| Morocco | 6 | 31.79 | 288.9 | 7 | 26.1 | 7.29 | 15.78 |
| Somalia | 6 | 5.15 | 148.8 | 5.1 | 8.3 | 3.04 | 6.88 |
| South Korea | 6 | 35.91 | 880.7 | 6.9 | 4.7 | 7.93 | 31.39 |
| Trinidad & Tobago | 6 | 10.69 | 432.1 | 11 | 18.6 | 9.05 | 25.94 |
| Belize | 5 | 17.19 | 274.9 | 17.1 | 24.1 | 5 | 10.71 |
| Chad | 5 | 15.45 | 128.9 | 6 | 6.1 | 2.81 | 5.67 |
| Lebanon | 5 | 33.85 | 593.7 | 11.2 | 33.7 | 5.08 | 17.58 |
| Slovakia | 5 | 48.67 | 803.3 | 6.5 | 20.5 | 13.38 | 30.2 |
| Tajikistan | 5 | 38.86 | 149.6 | 6.1 | 14.2 | 3.8 | 9.86 |
| Australia | 4 | -25.27 | 1849.8 | 5.6 | 29 | 9.27 | 27.23 |
| Georgia | 4 | 42.32 | 329.7 | 5.8 | 21.7 | 11 | 30.09 |
| Ghana | 4 | 7.95 | 228.5 | 2.5 | 10.9 | 3.81 | 9.65 |
| Malaysia | 4 | 4.21 | 337.2 | 16.7 | 15.6 | 6.99 | 15.71 |
| New Zealand | 4 | -40.90 | 1686.8 | 6.2 | 30.8 | 8.94 | 27.47 |
| Singapore | 4 | 1.35 | 703 | 5.5 | 6.1 | 6.91 | 21.47 |
| Tunisia | 4 | 33.89 | 256.1 | 8.5 | 26.9 | 7.35 | 18.98 |
| Burkina Faso | 3 | 12.24 | 158.1 | 7.3 | 5.6 | 2.95 | 6.73 |
| China | 3 | 35.86 | 377.6 | 9.2 | 6.2 | 7.86 | 24.42 |
| Costa Rica | 3 | 9.75 | 487.4 | 9.1 | 25.7 | 6.7 | 18.75 |
| Ivory Coast | 3 | 7.54 | 166.9 | 2.4 | 10.3 | 3.25 | 6.38 |
| Jamaica | 3 | 18.11 | 461.8 | 11.3 | 24.7 | 7.02 | 18.8 |
| Kenya | 3 | -0.02 | 302.4 | 3.1 | 7.1 | 3.42 | 7.08 |
| Libya | 3 | 26.34 | 254 | 10.2 | 32.5 | 5.74 | 9.56 |
| Niger | 3 | 17.61 | 89.1 | 2.4 | 5.5 | 2.58 | 5.85 |
| Nigeria | 3 | 9.08 | 183.2 | 3.1 | 8.9 | 2.98 | 7.43 |
| Benin | 2 | 9.31 | 198.1 | 1 | 9.6 | 2.92 | 5.54 |
| Guinea | 2 | 9.95 | 131 | 2.4 | 7.7 | 3.3 | 8.64 |
| Paraguay | 2 | -23.44 | 385.4 | 9.6 | 20.3 | 5.53 | 16.25 |
| Togo | 2 | 8.62 | 169.7 | 2.4 | 8.4 | 3.39 | 7.99 |
| Venezuela | 2 | 6.42 | 461.1 | 7 | 25.6 | 4.81 | 16.94 |
| Nepal | 1 | 28.39 | 171.5 | 7.2 | 4.1 | 4.46 | 12.33 |
| Zambia | 1 | -13.13 | 266.9 | 4.5 | 8.1 | 2.9 | 5.28 |
| Ethiopia | 0.9 | 9.15 | 155.7 | 4.3 | 4.5 | 2.85 | 7.8 |
| Jordan | 0.9 | 30.59 | 358.6 | 12.7 | 35.5 | 4.58 | 8.78 |
| Gambia | 0.8 | 13.44 | 74.8 | 1.9 | 10.3 | 3.34 | 8.09 |
| Thailand | 0.8 | 15.87 | 318.6 | 7 | 10 | 8.88 | 24.83 |
| Madagascar | 0.7 | -18.77 | 194.8 | 4.5 | 5.3 | 3.35 | 8.07 |
| Malawi | 0.7 | -13.25 | 277.7 | 4.5 | 5.8 | 3.47 | 5.66 |
| Uzbekistan | 0.7 | 41.38 | 182.4 | 6.5 | 16.6 | 4.94 | 14.5 |
| Sri Lanka | 0.5 | 7.87 | 214.4 | 10.7 | 5.2 | 6.63 | 21.11 |
| Syria | 0.5 | 34.80 | 298.5 | 13.5 | 27.8 | 5.72 | 9.87 |
| Zimbabwe | 0.5 | 29.15 | 1.8 | 15.5 | 3.59 | 8.59 | 4.52 |
| Botswana | 0.4 | -22.33 | 236.7 | 5.8 | 18.9 | 4.35 | 11.48 |
| Tanzania | 0.4 | -6.37 | 229.2 | 5.7 | 8.4 | 3.17 | 6.6 |
| Angola | 0.3 | -11.20 | 190.2 | 4.5 | 8.2 | 2.7 | 5.73 |
| Mozambique | 0.2 | -18.67 | 192.2 | 3.3 | 7.2 | 3.18 | 6.24 |
| Rwanda | 0.2 | -1.94 | 220 | 5.1 | 5.8 | 3.11 | 6.89 |
| Burundi | 0.08 | -3.37 | 194.8 | 5.1 | 5.4 | 3.07 | 7.23 |
| Bhutan | 0 | 27.51 | 137.1 | 10.3 | 6.4 | 3.99 | 12.99 |
| Cambodia | 0 | 12.57 | 198.9 | 6.4 | 3.9 | 4.25 | 11.03 |
| Eritrea | 0 | 15.18 | 142.6 | 5.1 | 5 | 2.85 | 7.8 |
| Fiji | 0 | -16.58 | 395 | 14.7 | 30.2 | 6.57 | 16.59 |
| Lao PDR | 0 | 19.86 | 255.6 | 6.4 | 5.3 | 4.24 | 9.86 |
| Lesotho | 0 | -29.61 | 199.2 | 4.5 | 16.6 | 4.18 | 10.58 |
| Mongolia | 0 | 46.86 | 308.7 | 4.7 | 20.6 | 5.19 | 12.82 |
| Namibia | 0 | -22.96 | 281.7 | 4.5 | 17.2 | 4.08 | 8.58 |
| Papua New Guinea | 0 | -6.31 | 342.8 | 17.9 | 21.3 | 3.98 | 10.47 |
| Timor-Leste | 0 | -8.87 | 155.2 | 6.7 | 3.8 | 4.19 | 9.28 |
| Uganda | 0 | 1.37 | 247.7 | 2.5 | 5.3 | 2.75 | 5.29 |
| Viet Nam | 0 | 14.06 | 279 | 6 | 2.1 | 5.36 | 16.46 |

